# Mendelian randomization of smoking behavior on cognitive status among older Americans

**DOI:** 10.1101/2019.12.11.19014522

**Authors:** Mingzhou Fu, Jessica D. Faul, Yuan Jin, Erin B. Ware, Kelly M. Bakulski

## Abstract

**Background:** Smoking is a likely risk factor for dementia, and smoking behavior has a strong genetic component. In this study, we jointly test the associations between cumulative genetic risk for smoking, smoking behavior, and cognitive status using a Mendelian randomization framework.

**Methods:** We conducted a cross-sectional study using the 2010 wave of the Health and Retirement Study database. Individuals aged between 50 and 90 were included. Smoking status was self-reported. Polygenic scores (PGSs) were calculated by weighting participant genotype by published smoking genome-wide association estimates. Cognitive status (normal, impaired, dementia) was measured using multiple assessments. A Mendelian randomization framework was used to infer causal relationships between smoking behavior and cognitive status via genetic instruments.

**Results:** Among European ancestry participants (N = 8,735), current smoking behavior was positively associated with cognitive impairment (OR = 1.62, 95% CI: 1.29, 2.01) relative to normal cognition. Using smoking PGS as an instrumental variable, a causal relationship was observed between current smoking and cognitive impairment (OR = 1.53, 95% CI: 1.07, 2.18). There were no associations between smoking PGS, smoking behaviors and cognitive status in the African ancestry study sample (N = 2,511).

**Conclusions:** Current smoking is a modifiable risk factor which causes cognitive impairment. Promotion of smoking cessation is important for public health. Further studies on dose and duration of smoking behaviors on cognitive impairment are critically needed, as well as in research other ancestries.

## Introduction

Dementia is a serious neurodegenerative disorder characterized by difficulties in a person’s daily life though memory loss, impaired language function, challenges in problem-solving, and changed cognitive status.^1^ In the United States (US), 14% of people over age 71 have dementia and the total medical care cost for all individuals with dementia was estimated at $277 billion in 2018.^2,3^ The most common type of dementia is Alzheimer’s disease (AD), which accounts for an estimated 60% to 80% of dementia cases.^4^ Given the large public health burden of dementia, understanding risk factors is extremely important.

Multiple environmental factors contribute to the incidence of dementia, such as internal environment factors like diabetes and obesity, as well as external exposures like environmental chemicals.^5,6^ Cigarette smoking is one of the most prevalent, but inconsistently, identified risk factors associated with dementia. In the US, with a prevalence of 15.5% among adults aged 18 years and older in 2016, smoking behavior contributed to 10.8% (95% Confidence Interval (CI): 3.0%, 19.8%) of the cases of dementia.^7,8^ However, the results of associations between cigarette smoking and dementia in a number of studies were contradictory.^9–11^ Further study of smoking as a risk factor for dementia and the characteristics that contribute to likelihood of smoking, such as genetics, are essential areas of investigation.

Smoking behavior is partially heritable. According to a former twin study, smoking initiation was 37% due to genetic factors.^12^ A previous genome-wide association study (GWAS) identified several genes associated with smoking behavior, including a synonymous 15q25 single nucleotide polymorphism (SNP) in the nicotinic receptor gene *CHRNA3* (rs1051730[A]), two 10q25 SNPs (rs1329650[G] and rs1028936[A]), and one 9q13 SNP in *EGLN2* (rs3733829[G]).^13^ A total of 566 genetic variants in 406 loci were recently detected in relation to smoking initiation, cessation, or heaviness.^14^ A single SNP, however, only accounts for a very small fraction of variability of the incidence of the disease or behavior.^15^ Researchers have proposed capturing the cumulative genetic risk across loci by using polygenic scores (PGSs). PGSs summarize an individual’s genetic association with a given trait into a single score and increase the amount of variation explained in a trait over single variants.^16,17^

The causal relationship between smoking behavior and cognitive impairment has not been rigorously assessed. Mendelian randomization is a method for genetic instrumental variable analyses, which can be applied to observational epidemiology studies.^18^ Because smoking behavior has a large genetic component, genetic sequences are established in early life, and child genotypes are a random sorting of parental genotypes, genetic predisposition to smoking can be used as an instrumental variable to infer causality testing with cognitive status. In the US nationally representative aging cohort, the Health and Retirement Study (HRS), we examined smoking behavior, genetic predisposition to smoking behavior, and cognitive status in European and African ancestry study samples. We first quantified the association between cumulative genetic risk for smoking and smoking behavior. Then we tested the association between smoking behavior and cognitive status. We next tested the assumption that cumulative genetic risk for smoking was not associated with cognitive status. Finally, we conducted a Mendelian randomization analysis to assess a potential causal relationship between smoking and cognitive status, using genetic risk for smoking as an instrumental variable.

## Methods

### 1. Health and Retirement Study

The HRS is a national longitudinal panel study of individuals over age 50 in the US.^16^ The HRS has collected data on health and economic information related to aging every two years since 1992. More than 43,000 individuals have participated to date.^19^ HRS data are publicly available (https://hrs.isr.umich.edu/). This cross-sectional analysis used the 2010 wave of HRS to maximize the number of cognitive impairment cases in a single wave. Asset and Health Dynamics among the Oldest Old (AHEAD) and Asset and Health Dynamics among the Oldest Old (CODA) are two sub-studies of HRS which included the oldest people in HRS.^20^ We excluded participants who were in these two studies as well as those younger than 50 or over 90 at 2010, because the diagnostic challenges, risk factors, and underlying neuropathological features of dementia are considerably different compared to octogenarians and younger.^21^ Our final study sample included 11,246 participants. (**Supplementary Figure 1**).

### 2. Cognitive status outcome assessment

The main outcome, cognitive status, was categorized in three levels as normal, cognitive impairment-non dementia (CIND), or dementia. The categorization method depended on whether the respondent could participate in the interview themselves, or due to physical or cognitive problems required a proxy respondent. Self-respondent categories were based on performance on a 27-point scale, including an immediate and delayed 10-noun free recall test, a serial 7 subtraction test, and a backward count from 20 test.^22^ The proxy categories were based on an 11-point scale. The proxy categorization used the proxy’s assessment of the respondent’s memory, whether the respondent had limitations in five instrumental activities of daily living (managing money, taking medication, preparing hot meals, using phones, and shopping for groceries), and whether the respondent had difficulty completing the interview because of a cognitive limitation. Cognitive status cut points were established by Crimmins et al. and have been validated clinically and empirically. The sensitivity of the cognitive outcome status variable was estimated as 78%.^23^

### 3. Exposure assessment and demographic characteristics

Smoking status was retrieved from the RAND HRS Longitudinal File.^24^ Participants were classified into never smokers, former smokers (reported ever smoking in the previous waves but not smoking at 2010), and current smokers (reported smoking at 2010).

Other demographic covariates used in the analysis included age, sex, years of education, rural/urban residence, body mass index (BMI), average exercise level, ever drinking alcohol, history of hypertension, diabetes, and/or depression, and stroke status. Age (yrs), sex (male/female), ever drinking alcohol and stroke history (Yes/No) were self-reported. Years of education was a continuous variable representing total number of years in school. Urban/rural residence was based on the 2003 Beale Rural-Urban Continuum Code, which divided living areas into urban, suburban, and ex-urban.^25^ BMI (kilograms/meters^2^) was generated by self-reported weight and height. Average exercise level was created from self-reported physical activity variables (vigorous, moderate, and mild) in the HRS. The original exercise variables were coded in a 5-point scale (1= every day to 5 = hardly ever or never). We reverse coded and averaged the three scores to represent an individual’s average exercise level. Hypertension, diabetes, or depression status (Yes/No) were generated from the self-report question of “Have you ever been told by doctor that you have … (the disease)?”. All variables were assessed at the 2010 HRS wave.

### 4. Genetic Data

Respondents provided saliva samples after reading and signing a consent form during an enhanced face-to-face interview in either 2006, 2008, 2010, or 2012 waves of the HRS. Details of the genotype collection and quality control are available elsewhere.^26^ Genotype measures were obtained using the Illumina HumanOmni2.5 BeadChip and genotyping was conducted by the Center for Inherited Disease Research. Genotype data that passed initial quality control were released to and analyzed by the Quality Assurance/Quality Control analysis team at the University of Washington.

A PGS is a single quantitative measure of genetic risk, which aggregates multiple individual loci across the human genome and weights them by effect sizes derived from the GWAS. The PGSs for AD and smoking initiation were downloaded from a published dataset by the HRS. The construction of PGSs for each phenotype was based on a single, large, replicated GWAS. The smoking PGS was created using results from a 2010 GWAS conducted by the Tobacco and Genetics Consortium.^13^ The AD PGS was created using results from a 2013 GWAS conducted by the International Genomics of Alzheimer’s Project.^27^ Apolipoprotein E (*APOE*) is an important independent risk gene for AD.^28^ We used the AD PGS without the two variants that contribute to *APOE* status (rs7412, rs429358), in addition to a binary variable of whether the individual is an *APOE*-*ε4* allele carrier in our fully adjusted model in the sensitivity analysis.^29^ *APOE*-*ε4* allele carrier was defined as individuals who have any copies of *ε4*.

Population stratification, that is, the allele frequency differences due to ancestry, could cause spurious associations between PGSs and the outcome of interest in disease studies.^30^ To control for confounding from population stratification and ancestry differences in genetic structures within populations, we conducted analyses separately by ancestry and adjusted for a set of five ancestry-specific principal components (PCs), as suggested by the HRS Documentation.^29^ Genetic ancestry (European or African) was identified through the union of PC analysis on genome-wide SNPs calculated across all participants and self-identified race/ethnicity.

### 5. Statistical Analysis

#### 5.1 Descriptive Analysis

All analyses were conducted in R version 3.5.1.^31^ The distributions of demographic, genetic, and behavioral factors were characterized using univariate analysis within included and excluded samples separately. Bivariate analyses were used to describe the distributions of covariates by outcome and exposure levels. For categorical variables, counts and frequencies for different cognitive or smoking levels were calculated. Chi-square tests were conducted to test for homogeneity between exposure or outcome groups. For continuous variables, means and standard deviations were calculated and a t-test or Wilcoxon rank sum test was used to assess differences between groups, as appropriate. Pearson correlations between each pair of variables were calculated and visualized using a heatmap. We further compared the smoking PGS and AD PGS using LD Hub to check for pleiotropy, defined as one gene which codes and controls the phenotype or expression of two or more different and unrelated traits.^32,33^ All data analyses were stratified by genetic ancestry.

#### 5.2 Regression Analysis

We used multivariable regression to test the associations between smoking PGS and smoking behavior on cognition status (**Supplementary Figure 2**). The cognition classification (normal, CIND, dementia) first suggested an ordinal analysis. We observed violations of the proportional odds assumption at all variables, except the smoking PGS. Thus, we elected to perform logistic regressions with the ***glm*()** function in the ***stats*** package for each level of cognition.^34^ We considered never smokers and those with normal cognition as the reference groups. In Model 1, we tested for an association between smoking PGS and smoking status. In Model 2, we tested for an association between smoking status and cognitive status. In Model 3, we tested for a direct association between smoking PGS and cognitive status, adjusting for smoking status. We conducted four sets of exposure/outcome comparisons in each model: A) former or never smokers with CIND or normal cognitive status; B) former or never smokers with dementia or normal cognitive status; C) current or never smokers with CIND or normal cognitive status; D) current or never smokers with dementia or normal cognitive status. We computed crude, unadjusted models. Our primary models were adjusted for age, sex, years of education, rural/urban residence, and five ancestry-specific PCs. Our primary analysis was in the European ancestry group. Population attributable fractions (PAFs) were also calculated for significant associations. We considered *P* value < .05 for statistical significance. We reported odds ratios (OR) and 95% CI.

#### 5.3 Mendelian Randomization Analysis

We used Mendelian randomization to examine whether the relationship between smoking behaviors and cognitive status is causal in our analysis. Our regression results met the three key assumptions of Mendelian randomization: 1) relevance assumption: the genetic variants associate with the risk factor of interest; 2) independence assumption: there are no unmeasured confounders of the association between genetic variants and outcome; 3) exclusion restriction: the genetic variants affect the outcome only through the effect on the risk factor of interest.^35^ Mendelian randomization was conducted in the same subsets (A-D) as the regression analyses. We used ***mr_input*()** and ***mr_ivw*()** functions from the ***MendelianRandomization*** package to build input and conduct estimation of causal effects.^36^ Within the function, we specified “Model 1” with smoking PGS as the exposure and smoking status as the outcome, and “Model 2” with smoking PGS as the exposure, and cognitive status as the outcome. The models were not adjusted as Mendelian randomization is not subject to confounding.^37^ ORs for the causal effect, along with their 95% CI and *P* value were reported.

#### 5.4 Stratified Analysis

We investigated whether the association between smoking PGS and cognitive status varied by smoking status. We tested for an association between smoking PGS and cognitive status, stratified by smoking status (never, former, and current), and visualized the association using scatter plots, with all the covariates set to the mean or reference group. We further tested for an interaction term between smoking PGS and smoking status on cognitive status.

#### 5.5 Sensitivity Analyses

We performed several sensitivity analyses to assess the robustness of our findings. First, we incorporated supplementary health risk factors. In addition to the covariates in the primary regression models, we adjusted for BMI, average exercise level, ever drinking, history of hypertension, diabetes, and/or depression, and stroke status. We next performed a set of sensitivity analyses including dementia genetic risk factors. In addition to the covariates in the primary regression models, we adjusted for AD PGS and any *APOE*-*ε4* allele carrier status.

To examine potential recall bias, defined as a systematic error resulting from differences in accuracy of recollections recalled by study participants,^38^ we compared the self-reported smoking statuses in 2008 (previous wave) to 2010 (primary analytic wave). We also compared the cognitive status in 2008 and 2010 to assess phenotype robustness.

Finally, all regression analyses were conducted in the African ancestry study sample. Code to produce all analyses in this manuscript are available (https://github.com/bakulskilab).

## Results

### 1. Descriptive analysis

Our primary analytic sample (N = 11,246) was 56.1% female, 77.7% European ancestry, with an average age of 65.0 years and 13.2 year of education. Participants included in our study sample did not differ from excluded samples with respect to hypertension, depression history or AD PGS (**Supplementary Table 1**). Our study sample had a lower mean age and education years, was more likely to be former smokers and be classified as having normal cognition than the excluded samples.

**Table 1.**
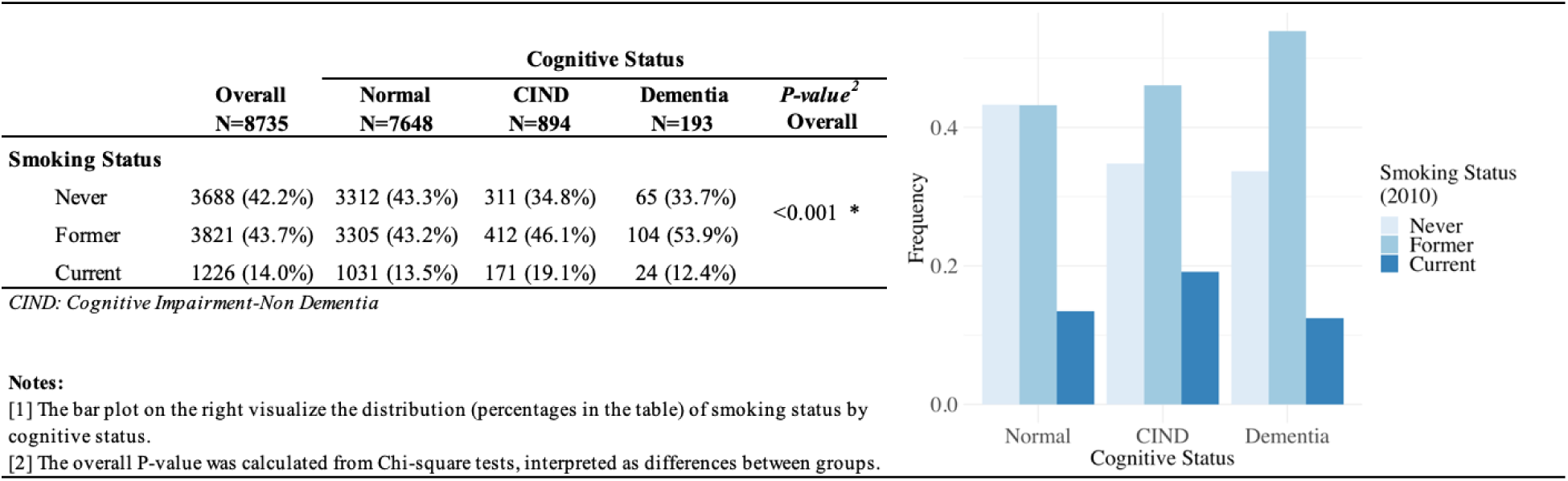
Cross tabulation of smoking status and cognitive status in European ancestry study sample^1^

In the European ancestry study sample (N = 8,735), cognitive status differed by smoking categories (*P* value < 0.001, **Table 1**). Current smoking was associated with higher proportion of CIND participants. Former smoking was positively associated with cognitive impairment.

Older age, lower educational attainment, rural residence, stroke, *APOE*-*ε4* allele carrier, higher AD PGS, lower BMI, never drinking, lower average exercise level, hypertension, diabetes, and depression were associated with cognitive impairment. Smoking PGS was not associated with cognitive status. Younger age, lower education, stroke, ever drinking, lower average exercise level, hypertension, diabetes, and depression were associated with higher levels of smoking behaviors. Higher smoking PGS was associated with former and current smoking behaviors (**Table 2**).

**Table 2.**
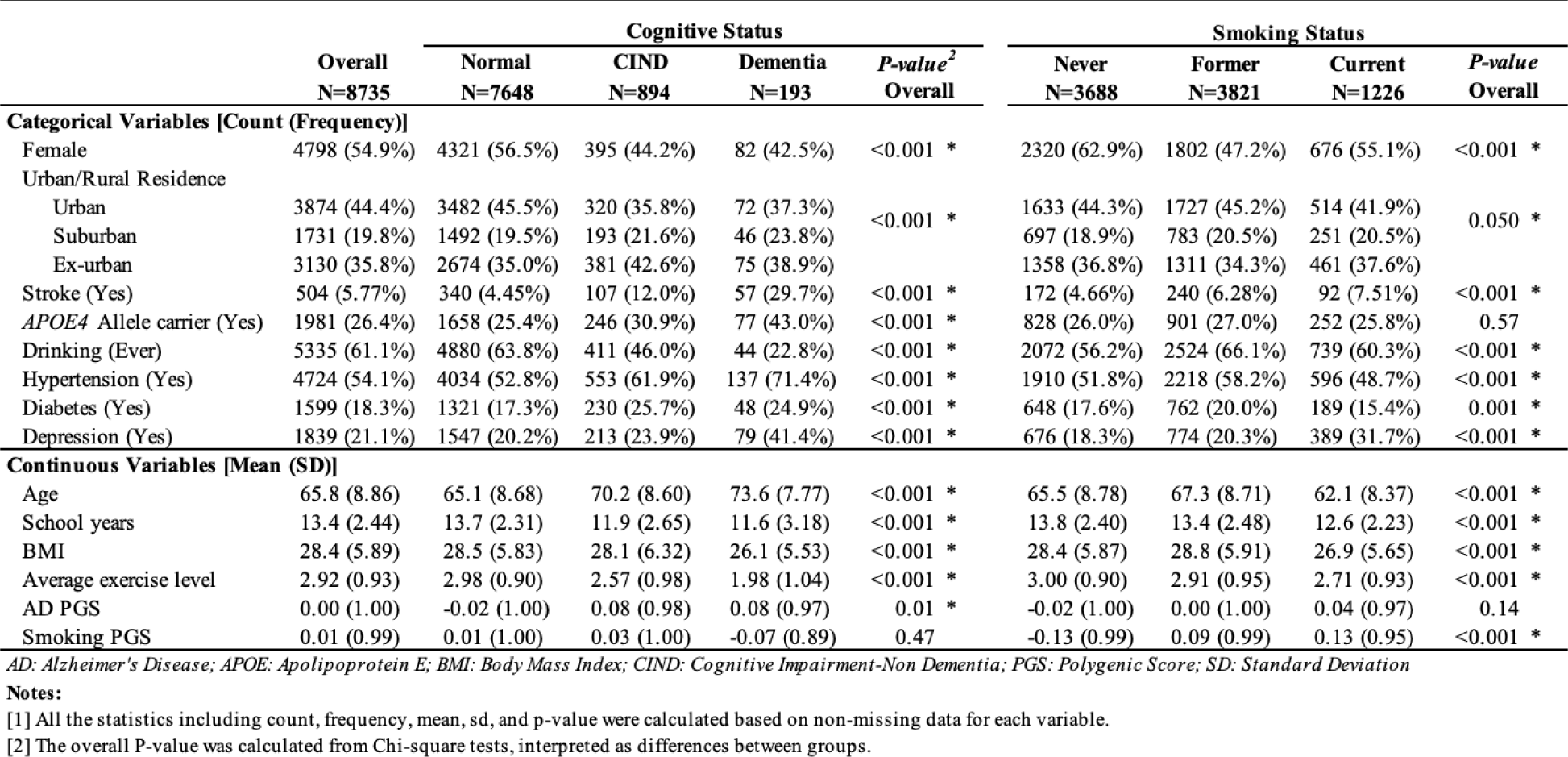
Bivariate characteristics stratified by cognitive status or smoking status in European ancestry study sample^1^

No correlation >|0.4| was observed between any two covariates (**Supplementary Figure 3**). Specifically, there were minor correlations observed between AD PGS and smoking PGS (r = 0.04, p < 0.01), and between AD PGS and smoking status (r = 0.02, p = 0.07). Those findings were consistent with results provided by LD Hub (r = 0.101, p = 0.649).^39^

### 2. Regression analyses

In the European ancestry sample, smoking PGS is highly associated with smoking status **(Table 3, Model 1)**. In the primary adjusted model, a one standard deviation unit increase in smoking PGS was associated with 1.27-1.28 times higher odds of former or current smoking behavior relative to never smoking. These findings were robust to additional adjustment for health status and AD genetics (**Supplemental Table 2**). PAF results suggested that 19.74% (95%CI: 11.03%, 28.46%) of the current smoking cases were attributed to smoking genetics, after adjusting for age, sex, years of education, rural/urban residence, and five ancestry-specific PCs (**Supplemental Table 3**).

**Table 3.** Results for logistic regression analyses among European ancestry samples^1^

|  | Former vs. Never |  | Current vs. Never |  |
| --- | --- | --- | --- | --- |
|  | OR (95 CI%)<sup>2</sup> | P value<sup>3</sup> | OR (95 CI%) | P value |
| **CIND vs. Normal** | **N = 7,340** |  | **N = 4,825** |  |
| **Model 1: Smoking Status ~ Smoking PGS** |  |  |  |  |
| Crude | 1.26 (1.20, 1.32) | <0.001 * | 1.30 (1.22, 1.39) | <0.001 * |
| Adjusted (demographic)<sup>4</sup> | 1.27 (1.20, 1.34) | <0.001 * | 1.28 (1.18, 1.38) | <0.001 * |
| **Model 2: Cognitive Status ~ Smoking Status** |  |  |  |  |
| Crude | 1.33 (1.14, 1.55) | <0.001 * | 1.77 (1.44, 2.15) | 0.001 * |
| Adjusted (demographic) | 0.98 (0.83, 1.16) | 0.63 | 1.62 (1.29, 2.01) | <0.001 * |
| **Model 3: Cognitive Status ~ Smoking PGS + Smoking Status (Direct effect of Smoking PGS)** |  |  |  |  |
| Crude | 0.98 (0.90, 1.06) | 0.45 | 1.09 (0.99, 1.20) | 0.20 |
| Adjusted (demographic) | 0.96 (0.88, 1.05) | 0.57 | 1.06 (0.95, 1.19) | 0.58 |
| **Dementia vs. Normal** | **N = 6,786** |  | **N = 4,432** |  |
| **Model 1: Smoking Status ~ Smoking PGS** |  |  |  |  |
| Crude | 1.27 (1.21, 1.34) | <0.001 * | 1.29 (1.20, 1.39) | <0.001 * |
| Adjusted (demographic) | 1.28 (1.21, 1.36) | <0.001 * | 1.27 (1.17, 1.38) | <0.001 * |
| **Model 2: Cognitive Status ~ Smoking Status** |  |  |  |  |
| Crude | 1.60 (1.18, 2.20) | 0.24 | 1.19 (0.73, 1.88) | 0.06 |
| Adjusted (demographic) | 1.12 (0.80, 1.57) | 0.86 | 1.12 (0.66, 1.85) | 0.24 |
| **Model 3: Cognitive Status ~ Smoking PGS + Smoking Status (Direct effect of Smoking PGS)** |  |  |  |  |
| Crude | 0.89 (0.77, 1.04) | 0.48 | 1.01 (0.81, 1.25) | 0.26 |
| Adjusted (demographic) | 0.89 (0.74, 1.07) | 0.52 | 1.00 (0.78, 1.28) | 0.68 |

CI: Confidence Interval; CIND: Cognitive Impairment-Non Dementia; OR: Odds Ratio; PC: Principal Component;
**Notes:**
[1] All the logistic regression analyses used "Never Smoking" and "Normal Cognitive Status" as references.
[2] The OR(95%CI) values for the first variable after the tilde (~) in each model.
[3] P value represents the statistical significance of the according coefficient(s).
[4] Adjusted models were adjusted for age, sex, urban/rural living, years of education and five ancestry-specific PC

Smoking behavior was associated with cognitive status in the European ancestry sample (**Table 3, Model 2**). In the primary adjusted model, current smokers relative to never smokers had 1.62 (95%CI: 1.29, 2.01) times odds of CIND relative to normal cognition. 11.35% (95%CI: 5.87%, 16.83%) of the CIND cases were attributed to current smoking, according to PAF analysis (**Supplemental Table 3**). After additional adjustment for health status, current smokers relative to never smokers had 1.38 (95%CI: 1.09, 1.75) times odds of CIND relative to normal cognition. These positive associations were even higher when additionally adjusted for AD genetics (OR = 1.64, 95% CI: 1.29, 2.08) (**Supplemental Table 2**). No associations were observed between current smoking and dementia, or between former smoking with either CIND or dementia.

Smoking PGS was not associated with cognitive status directly (**Table 3, Model 3**). This reflects an important assumption of Mendelian randomization, that the instrumental variable (smoking PGS) is only associated with the outcome through the primary exposure and not independently.

### 3. Mendelian randomization analyses

We observed strong positive association between smoking PGS and smoking status, positive association between smoking behavior and cognitive status, and no association between smoking PGS and cognitive status, which jointly met the key assumptions of relevance, independence, and exclusion restriction for Mendelian randomization. A test of inferred causality of smoking behavior on cognitive impairment was conducted using smoking PGS as the instrumental variable. According to **Table 4**, current smokers relative to never smokers had 1.53 (95%CI: 1.07, 2.18) times risk of CIND relative to normal cognition. However, no significant causal effect was found for current smoking on dementia, or former smoking on either CIND or dementia. Current smokers relative to never smokers had 1.07 (95%CI: 0.47, 2.45) times risk of dementia relative to normal cognition. Former smokers relative to never smokers had 0.97 (95%CI: 0.70, 1.36) times risk of CIND and 0.71 (95%CI: 0.37, 1.34) times risk of dementia relative to normal cognition. Taken as a whole, there is a causal relationship only between current smoking and cognitive impairment.

**Table 4.**
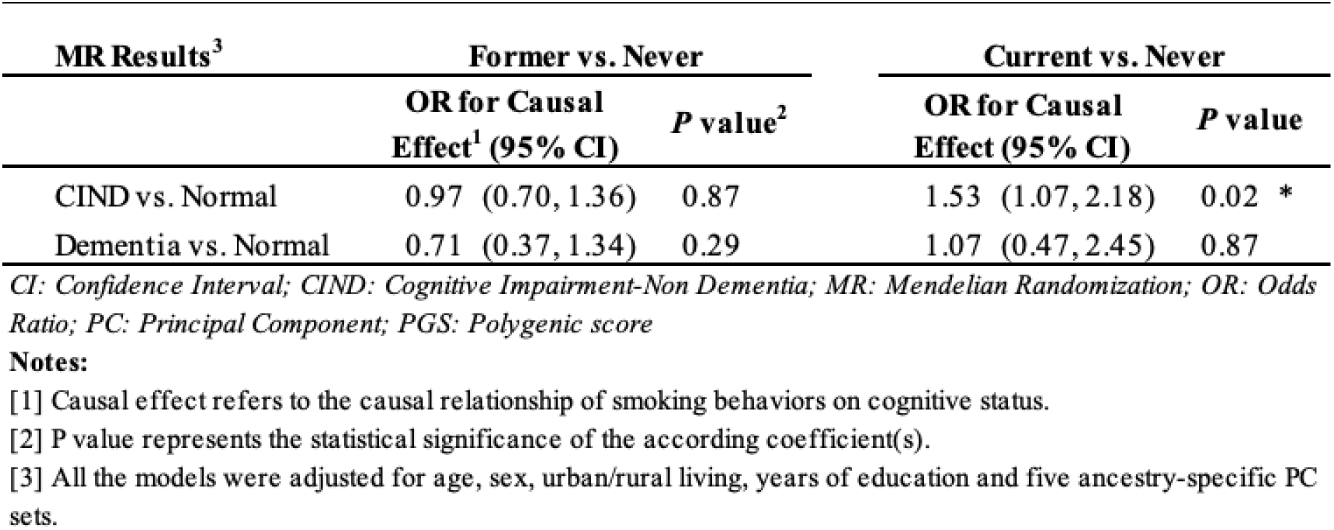
Mendelian Randomization results in European ancestry

| MR Results<sup>3</sup> | Former vs. Never |  | Current vs. Never |  |
| --- | --- | --- | --- | --- |
|  | OR for Causal Effect<sup>1</sup> (95% CI) | P value<sup>2</sup> | OR for Causal Effect (95% CI) | P value |
| CIND vs. Normal | 0.97 (0.70, 1.36) | 0.87 | 1.53 (1.07, 2.18) | 0.02 * |
| Dementia vs. Normal | 0.71 (0.37, 1.34) | 0.29 | 1.07 (0.47, 2.45) | 0.87 |

CI: Confidence Interval; CIND: Cognitive Impairment-Non Dementia; MR: Mendelian Randomization; OR: Odds Ratio; PC: Principal Component; PGS: Polygenic score
**Notes:**
[1] Causal effect refers to the causal relationship of smoking behaviors on cognitive status.
[2] P value represents the statistical significance of the according coefficient(s).
[3] All the models were adjusted for age, sex, urban/rural living, years of education and five ancestry-specific PC sets.

### 4. Stratified analysis

We did not observe an association between smoking PGS and cognitive status in the European ancestry group, in any of the smoking strata (never, former, current) **(Table 5)**. Regardless of statistical significance, current and never smokers with higher smoking PGS had higher probability of CIND or dementia versus normal status. We observed a significant interaction between former smoking status and smoking PGS on CIND cognitive status (β = −0.200, p value = 0.02).

**Table 5.** Results for stratified analyses on association between smoking PGS and cognitive status in European ancestry sample^1^

| Stratified analysis |  | CIND vs. Normal |  |  | Dementia vs. Normal |  |  |
| --- | --- | --- | --- | --- | --- | --- | --- |
| Smoker Group |  | N | OR for Smoking PGS (95 CI%) | P value<sup>2</sup> | N | OR for Smoking PGS (95 CI%) | P value |
| Never | Crude | 3,623 | 1.07 (0.95, 1.20) | 0.26 | 3,377 | 1.01 (0.79, 1.29) | 0.94 |
|  | Adjusted (demographic)<sup>3</sup> | 3,623 | 1.06 (0.93, 1.21) | 0.41 | 3,377 | 1.00 (0.75, 1.34) | 0.97 |
| Former | Crude | 3,717 | 0.91 (0.82, 1.01) | 0.08 | 3,409 | 0.83 (0.68, 1.01) | 0.06 |
|  | Adjusted (demographic) | 3,717 | 0.88 (0.78, 1.00) | 0.05 * | 3,409 | 0.81 (0.64, 1.03) | 0.09 |
| Current | Crude | 1,202 | 1.13 (0.95, 1.33) | 0.17 | 1,055 | 1.01 (0.66, 1.54) | 0.97 |
|  | Adjusted (demographic) | 1,202 | 1.07 (0.88, 1.30) | 0.52 | 1,055 | 1.00 (0.60, 1.67) | 0.99 |
| **Test for Interactions between Smoking Status and Smoking PGS** |  |  |  |  |  |  |  |
|  |  | N | β Coefficient for Interaction Term | P value | N | β Coefficient for Interaction Term | P value |
| Former vs. Never | Crude | 7,340 | -0.160 | 0.04 * | 6,786 | -0.201 | 0.21 |
|  | Adjusted (demographic) | 7,340 | -0.200 | 0.02 * | 6,786 | -0.271 | 0.10 |
| Current vs. Never | Crude | 4,825 | 0.051 | 0.63 | 4,432 | 0.000 | 1.00 |
|  | Adjusted (demographic) | 4,825 | 0.004 | 0.97 | 4,432 | -0.042 | 0.87 |

CI: Confidence Interval; CIND: Cognitive Impairment-Non Dementia; OR: Odds Ratio; PC: Principal Component; PGS: Polygenic Score
**Notes:**
[1] All the analyses used "Normal Cognitive Status" as references and were stratified by smoking status
[2] P value represents the statistical significance of the according coefficient(s).
[3] Adjusted models were adjusted for age, sex, urban/rural living, years of education and five ancestry-specific PC sets.

### 5. Sensitivity analyses

We assessed consistency in response to smoking questionnaires. There was no observation reported as never smoker in the 2010 study sample who reported former or current smoker status in 2008, indicating lack of smoking misclassification. We further assessed sensitivity of cognitive status measures over time. In 2008, there were 318 (4.2%) CIND and 9 (0.1%) dementia observations that had normal cognitive status in the 2010 study sample **(Supplemental Table 4)**.

The African ancestry analytic sample included 2,511 participants. We combined the CIND and dementia groups to increase the power of the analysis. No associations were observed between smoking PGS and smoking status, smoking PGS and cognitive status, or smoking status and cognitive status **(Supplemental Table 5)**. The regression results did not meet the Mendelian randomization assumptions, thus, we did not pursue further causal inference testing.

## Discussion

The present cross-sectional study was conducted among older adults from the HRS wave 2010. To our knowledge, this is the first study using Mendelian randomization framework to examine the relationship between smoking behaviors and cognitive status with a genetic instrument of cumulative genetic risk for smoking behavior. In the European ancestry individuals, after adjusting for age, sex, years of education, rural/urban residence and five ancestry-specific PCs, we found a strong positive association between smoking PGS and smoking behaviors – one standard deviation increase in smoking PGS was associated with 1.27-1.28 times odds of former or current smoking behavior relative to never smoking. We also observed a positive association between current smoking and CIND relative to normal cognition: current smokers had 1.62 times odds of CIND (95%CI: 1.29, 2.01) relative to never smokers. No association was found between smoking PGS and cognitive status. A significant causal relationship was found between current smoking and CIND.

Smoking PGS was positively associated with smoking behaviors in the European ancestry sample only. The GWAS of smoking behaviors conducted by Tobacco and Genetics Consortium in 2010 focused exclusively on participants of the European ancestry.^13^ Given that our methods for computing the smoking PGS depended on summary statistics from this GWAS, risk alleles identified from this study may be specific to the European ancestry and result in limited generalizability to other ancestral groups.^40^ Furthermore, only 2,511 individuals were included in our African ancestry analysis, versus 8,735 in the European ancestry analysis. Thus, with the sample size and the European ancestry reference GWAS, we were not surprised by the null association between smoking PGS and smoking behaviors observed in the African ancestry sample.

We found current smokers compared with never smokers were at increased risk of CIND in the European ancestry sample, which was consistent with previous studies.^41–43^ Potential biological mechanism for this relationship could implicate elevated chronic oxidative stress in the brain and other organ systems of smokers which may trigger the AD-pathophysiological process. Computed tomography and magnetic resonance based studies also found supportive evidence of abnormalities in brain morphology, perfusion and neurochemistry in smokers.^44^ Our null associations between former smoking behavior and cognitive impairment were also similar to some previous studies.^45,46^ In addition to examining the association, we also tested the causal relationship between smoking behaviors and cognitive status in the European ancestry using Mendelian randomization. This method, which limits the possibility of confounding, and the using of multiple variants (PGS) increased the power and test assumptions.^18^

Mortality selection is a concern for both dementia and smoking. Smokers may die prematurely from other smoking-related diseases before developing dementia. According to a study on HRS samples from 1992-2005, the genotyped HRS respondents were longer-lived as compared with their non-genotyped respondents.^47^ Thus, the analysis of smoking and dementia was subject to survival bias; that is, the samples in the study were biased toward healthier smokers – individuals who survived or did not experience significant smoking-related morbidity.^48,49^ Furthermore, in reference to **Supplementary Table 1**, our study sample captured a lower proportion of dementia (included vs. excluded: 3.22% vs. 10.2%) individuals. And the development of other smoking-related morbidity, such as cardiovascular diseases and cancer, may also impede participation in the longitudinal HRS study. These could result in a selection bias in that the study sample is not representative of the smokers in the general older population. Both the survival and selection bias will bias the association between smoking behaviors and cognitive impairment toward the null. Therefore, the smoking-related risk for cognitive impairment was likely underestimated in our study.

In our current study, smoking status was categorized cross-sectionally using retrospective assessments, and information on the number of cigarettes consumed or duration of smoking or cessation was not included. Further studies, with more expansive smoking variables, will be needed to test the dose effect of smoking on cognitive status. A similar study should be replicated in an African ancestry with a larger sample size and PGSs based on GWASs among the African ancestry. These results could also be strengthened by a longitudinal assessment of incident dementia.

Unlike genetic factors, smoking behaviors are modifiable, which can be effectively altered, and promote a significant decrease in cases of dementia. The significant causal relationship found in our study provide evidence to support current smoking as a risk factor for cognitive impairment in later life. The null effect of former smoking suggests, while preliminary, a plausible protective effect of smoking cessation. Thus, promotion of smoking cessation could be the most effective strategy for lowering dementia prevalence over time at the population level.

In conclusion, in a cross-sectional analysis of the HRS, we found evidence of causality of current smoking as a risk factor on cognitive impairment using genetic instruments. Additional studies on dose and time-span effect of smoking behaviors on cognitive impairment are critically needed, as well as studies in other ancestries. Public health campaigns should make explicit the connection between current smoking and dementia as yet another reason to quit.

## Data Availability

Data used in this manuscript are publicly available:
https://hrs.isr.umich.edu/data-products
Code used to produce analyses are publicly available: 
https://github.com/bakulskilab

## Acknowledgments

All authors were supported by a grant from the National Institute on Aging (R01 AG055406). Dr. Bakulski was supported by grants from the National Institute for Environmental Health Sciences and National Institute for Minority Health and Health Disparities (R01 ES025531; R01 ES025574; OD023285; and R01 MD013299). Dr. Ware was supported by grants from the National Institute for Minority Health and Health Disparities (R01 MD012145) and the National Institute on Aging (R01 AG055654). This work was supported by the University of Michigan, Michigan Lifestage Environmental Exposures and Disease (M-LEEaD) National Institute for Environmental Health Sciences Core Center (P30 ES017885).

## Author Contributions

Conceived and designed the analysis plan: MF, KB, EW. Managed the dataset: MF. Analyzed the data: MF. Shadow analysis and code review: YJ. Contributed to the interpretation: MF, KB, EW, JD. Writing and editing of the manuscript: MF, KB, EW, JD.

## Acronyms

Alzheimer’s disease (AD), Apolipoprotein E (APOE), body mass index (BMI), confidence interval (CI), genome wide association study (GWAS), Health and Retirement Study (HRS), odds ratio (OR), polygenic score (PGS), single nucleotide polymorphism (SNP).

## Notes

**Declaration of competing financial interests** The authors declare they have no actual or potential competing financial interests.

### Competing Interest Statement

The authors have declared no competing interest.

## References

1. McKhann GM, Knopman DS, Chertkow H, et al. The diagnosis of dementia due to Alzheimer’s disease: recommendations from the National Institute on Aging-Alzheimer’s Association workgroups on diagnostic guidelines for Alzheimer’s disease. Alzheimers Dement. 2011;7(3):263–269. doi:10.1016/j.jalz.2011.03.005

2. Plassman BL, Langa KM, Fisher GG, et al. Prevalence of dementia in the United States: the aging, demographics, and memory study. Neuroepidemiology. 2007;29(1-2):125–132. doi:10.1159/000109998

3. Wilson RS, Segawa E, Boyle PA, Anagnos SE, Hizel LP, Bennett DA. The natural history of cognitive decline in Alzheimer’s disease. Psychol Aging. 2012;27(4):1008–1017. doi:10.1037/a0029857

4. Barker WW, Luis CA, Kashuba A, et al. Relative frequencies of Alzheimer disease, Lewy body, vascular and frontotemporal dementia, and hippocampal sclerosis in the State of Florida Brain Bank. Alzheimer Dis Assoc Disord. 2002;16(4):203–212.

5. Jellinger KA. The enigma of mixed dementia. Alzheimers Dement. 2007;3(1):40–53. doi:10.1016/j.jalz.2006.09.002

6. Chin-Chan M, Navarro-Yepes J, Quintanilla-Vega B. Environmental pollutants as risk factors for neurodegenerative disorders: Alzheimer and Parkinson diseases. Front Cell Neurosci. 2015;9:124. doi:10.3389/fncel.2015.00124

7. Jamal A, Phillips E, Gentzke AS, et al. Current Cigarette Smoking Among Adults - United States, 2016. MMWR Morb Mortal Wkly Rep. 2018;67(2):53–59. doi:10.15585/mmwr.mm6702a1

8. Barnes DE, Yaffe K. The projected effect of risk factor reduction on Alzheimer’s disease prevalence. Lancet Neurol. 2011;10(9):819–828. doi:10.1016/S1474-4422(11)70072-2

9. Lee PN. Smoking and Alzheimer’s disease: a review of the epidemiological evidence. Neuroepidemiology. 1994;13(4):131–144. doi:10.1159/000110372

10. Van Duijn CM, Clayton DG, Chandra V, et al. Interaction between genetic and environmental risk factors for Alzheimer’s disease: a reanalysis of case-control studies. Genet Epidemiol. 1994;11(6):539–551. doi:10.1002/gepi.1370110609

11. Anstey KJ, von Sanden C, Salim A, O’Kearney R. Smoking as a risk factor for dementia and cognitive decline: a meta-analysis of prospective studies. Am J Epidemiol. 2007;166(4):367–378. doi:10.1093/aje/kwm116

12. Seglem KB, Waaktaar T, Ask H, Torgersen S. Genetic and environmental influences on adolescents’ smoking involvement: a multi-informant twin study. Behav Genet. 2015;45(2):171–180. doi:10.1007/s10519-015-9706-x

13. Tobacco and Genetics Consortium. Genome-wide meta-analyses identify multiple loci associated with smoking behavior. Nat Genet. 2010;42(5):441–447. doi:10.1038/ng.571

14. Liu M, Jiang Y, Wedow R, et al. Association studies of up to 1.2 million individuals yield new insights into the genetic etiology of tobacco and alcohol use. Nat Genet. 2019;51(2):237–244. doi:10.1038/s41588-018-0307-5

15. Justice AE, Winkler TW, Feitosa MF, et al. Genome-wide meta-analysis of 241,258 adults accounting for smoking behaviour identifies novel loci for obesity traits. Nat Commun. 2017;8:14977. doi:10.1038/ncomms14977

16. Ware EB, Schmitz LL, Faul J, et al. Heterogeneity in polygenic scores for common human traits. bioRxiv. February 2017:106062. doi:10.1101/106062

17. Escott-Price V, Sims R, Bannister C, et al. Common polygenic variation enhances risk prediction for Alzheimer’s disease. Brain. 2015;138(Pt 12):3673–3684. doi:10.1093/brain/awv268

18. Davey Smith G, Hemani G. Mendelian randomization: genetic anchors for causal inference in epidemiological studies. Hum Mol Genet. 2014;23(R1):R89–98. doi:10.1093/hmg/ddu328

19. Sonnega A, Faul JD, Ofstedal MB, Langa KM, Phillips JWR, Weir DR. Cohort Profile: the Health and Retirement Study (HRS). Int J Epidemiol. 2014;43(2):576–585. doi:10.1093/ije/dyu067

20. Health and Retirement Study (HRS) & Asset and Health Dynamics among the Oldest Old (AHEAD) | sgim.org. https://www.sgim.org/communities/research/dataset-compendium/health-and-retirement-study-hrs--asset-and-health-dynamics-among-the-oldest-old-ahead. Accessed October 24, 2019.

21. Bullain SS, Corrada MM. Dementia in the oldest old. Continuum (Minneap Minn). 2013;19(2 Dementia):457–469. doi:10.1212/01.CON.0000429172.27815.3f

22. Ofstedal MB, Fisher G. Documentation of Cognitive Functioning Measures in the Health and Retirement Study. Institute for Social Research, University of Michigan; 2005. doi:10.7826/ISR-UM.06.585031.001.05.0010.2005

23. Crimmins EM, Kim JK, Langa KM, Weir DR. Assessment of Cognition Using Surveys and Neuropsychological Assessment: The Health and Retirement Study and the Aging, Demographics, and Memory Study. J Gerontol B Psychol Sci Soc Sci. 2011;66B(Suppl 1):i162–i171. doi:10.1093/geronb/gbr048

24. Monica 1776 Main Street Santa, California 90401-3208. RAND HRS Longitudinal File 2016 (V1). https://www.rand.org/well-being/social-and-behavioral-policy/centers/aging/dataprod/hrs-data.html. Accessed October 22, 2019.

25. USDA ERS - Rural-Urban Continuum Codes. https://www.ers.usda.gov/data-products/rural-urban-continuum-codes/. Accessed October 22, 2019.

26. David R. W. Quality Control Report for Genotypic Data. September 2013. http://hrsonline.isr.umich.edu/sitedocs/genetics/HRS2_qc_report_SEPT2013.pdf?_ga=2.160262008.498457094.1571249480-1101446680.1569101058.

27. Lambert JC, Ibrahim-Verbaas CA, Harold D, et al. Meta-analysis of 74,046 individuals identifies 11 new susceptibility loci for Alzheimer’s disease. Nat Genet. 2013;45(12):1452–1458. doi:10.1038/ng.2802

28. Heffernan AL, Chidgey C, Peng P, Masters CL, Roberts BR. The Neurobiology and Age-Related Prevalence of the ε4 Allele of Apolipoprotein E in Alzheimer’s Disease Cohorts. J Mol Neurosci. 2016;60(3):316–324. doi:10.1007/s12031-016-0804-x

29. HRS Polygenic Scores 2006-2012 Genetic Data - Release 3 | Health and Retirement Study. https://hrs.isr.umich.edu/news/hrs-polygenic-scores-2006-2012-genetic-data-release-3. Accessed October 18, 2019.

30. Price AL, Patterson NJ, Plenge RM, Weinblatt ME, Shadick NA, Reich D. Principal components analysis corrects for stratification in genome-wide association studies. Nat Genet. 2006;38(8):904–909. doi:10.1038/ng1847

31. R: The R Project for Statistical Computing. https://www.r-project.org/. Accessed October 18, 2019.

32. Solovieff N, Cotsapas C, Lee PH, Purcell SM, Smoller JW. Pleiotropy in complex traits: challenges and strategies. Nat Rev Genet. 2013;14(7):483–495. doi:10.1038/nrg3461

33. Bulik-Sullivan B, Finucane HK, Anttila V, et al. An atlas of genetic correlations across human diseases and traits. Nat Genet. 2015;47(11):1236–1241. doi:10.1038/ng.3406

34. glm function | R Documentation. https://www.rdocumentation.org/packages/stats/versions/3.6.1/topics/glm. Accessed October 19, 2019.

35. Davies NM, Holmes MV, Davey Smith G. Reading Mendelian randomisation studies: a guide, glossary, and checklist for clinicians. BMJ. 2018;362:k601. doi:10.1136/bmj.k601

36. Olena Y. Mendelian Ramdomization. https://www.ncbi.nlm.nih.gov/projects/gap/cgi-bin/study.cgi?study_id=phs000428.v2.p2. Accessed October 19, 2019.

37. Smith GD, Ebrahim S. “Mendelian randomization”: can genetic epidemiology contribute to understanding environmental determinants of disease? Int J Epidemiol. 2003;32(1):1–22. doi:10.1093/ije/dyg070

38. Coughlin SS. Recall bias in epidemiologic studies. J Clin Epidemiol. 1990;43(1):87–91. doi:10.1016/0895-4356(90)90060-3

39. Sullivan B. Lookup Center: Lookup existing LD score regression analysis results. http://ldsc.broadinstitute.org/. Published 2015. Accessed October 18, 2019.

40. Martin AR, Gignoux CR, Walters RK, et al. Human Demographic History Impacts Genetic Risk Prediction across Diverse Populations. Am J Hum Genet. 2017;100(4):635–649. doi:10.1016/j.ajhg.2017.03.004

41. Aggarwal NT, Bienias JL, Bennett DA, et al. The relation of cigarette smoking to incident Alzheimer’s disease in a biracial urban community population. Neuroepidemiology. 2006;26(3):140–146. doi:10.1159/000091654

42. Juan D, Zhou DHD, Li J, Wang JYJ, Gao C, Chen M. A 2-year follow-up study of cigarette smoking and risk of dementia. Eur J Neurol. 2004;11(4):277–282. doi:10.1046/j.1468-1331.2003.00779.x

43. Ott A, Slooter AJ, Hofman A, et al. Smoking and risk of dementia and Alzheimer’s disease in a population-based cohort study: the Rotterdam Study. Lancet. 1998;351(9119):1840–1843. doi:10.1016/s0140-6736(97)07541-7

44. Durazzo TC, Meyerhoff DJ, Nixon SJ. Chronic cigarette smoking: implications for neurocognition and brain neurobiology. Int J Environ Res Public Health. 2010;7(10):3760–3791. doi:10.3390/ijerph7103760

45. Reitz C, den Heijer T, van Duijn C, Hofman A, Breteler MMB. Relation between smoking and risk of dementia and Alzheimer disease: the Rotterdam Study. Neurology. 2007;69(10):998–1005. doi:10.1212/01.wnl.0000271395.29695.9a

46. Merchant C, Tang MX, Albert S, Manly J, Stern Y, Mayeux R. The influence of smoking on the risk of Alzheimer’s disease. Neurology. 1999;52(7):1408–1412. doi:10.1212/wnl.52.7.1408

47. Domingue BW, Belsky DW, Harrati A, Conley D, Weir DR, Boardman JD. Mortality selection in a genetic sample and implications for association studies. Int J Epidemiol. 2017;46(4):1285–1294. doi:10.1093/ije/dyx041

48. Riggs JE. Smoking and Alzheimer’s disease: protective effect or differential survival bias? Lancet. 1993;342(8874):793–794. doi:10.1016/0140-6736(93)91547-y

49. Mayeda ER, Tchetgen Tchetgen EJ, Power MC, et al. A Simulation Platform for Quantifying Survival Bias: An Application to Research on Determinants of Cognitive Decline. Am J Epidemiol. 2016;184(5):378–387. doi:10.1093/aje/kwv451

